# Steroidome Dysregulation and Complement C4 Copy Number Variation in Men with Central Serous Chorioretinopathy

**DOI:** 10.1101/2025.10.19.25338305

**Authors:** Marta Zola, Ji Hoon Han, Mathieu Quinodoz, Elodie Bousquet, Alexandre Matet, Laura Kowalczuk, Alejandra Daruich, Min Zhao, Mukhtar Ullah, Carlo Rivolta, Chiara Eandi, Yvan Arsenijevicz, Eric Pussard, Francine Behar-Cohen

**Affiliations:** Centre de Recherche des Cordeliers, INSERM, Université Paris Cité, Sorbonne Université, Physiopathology of ocular diseases: Therapeutic innovations, Paris, France; Ophthalmopole Cochin University Hospital, Assistance Publique-Hôpitaux de Paris, France; Institute of Molecular and Clinical Ophthalmology Basel (IOB), Basel, Switzerland; Department of Ophthalmology, University Hospital Basel, Basel, Switzerland; Department of Genetics and Genome Biology, University of Leicester, Leicester, UK; Institut Curie, Department of Ophthalmology, Paris, France; Jules-Gonin Eye Hospital, Fondation Asile des aveugles, Lausanne, Switzerland; Faculty of Biology and Medicine, University of Lausanne, Lausanne, Switzerland; Université Paris Cité, Paris, France; Department of Ophthalmology, Necker Enfants Malades University Hospital, Assistance Publique-Hôpitaux de Paris, France; Department of Genetic and Hormonology, Bicêtre Hospital, Assistance Publique-Hôpitaux de Paris, University of Paris-Saclay, Le Kremlin Bicêtre, France; Hopital Foch, Suresnes, France

## Abstract

**Background:** Central serous chorioretinopathy (CSCR) predominantly affects middle-aged men, in which systemic glucocorticoid exposure is a well-established risk factor. Genetic studies have implicated copy number variation (CNV) in complement component 4 (C4), located within the RCCX module alongside the steroid 21-hydroxylase gene (CYP21A2), in modifying disease susceptibility. However, the relationship between adrenal steroid metabolism and C4 CNV in CSCR has not been investigated.

**Methods:** We analyzed the serum steroidome of 45 male CSCR patients and 49 age-matched healthy male controls using liquid chromatography–tandem mass spectrometry. In a subset of 20 patients with chronic CSCR, genomic C4A and C4B copy numbers were quantified by qPCR and correlated with circulating steroid metabolites, including indices of CYP21A2 activity.

**Results:** CSCR patients exhibited significant alterations in steroid metabolism compared with controls. In the glucocorticoid pathway, 17-hydroxyprogesterone (17-OHP) was elevated, cortisone was reduced, and the 11-deoxycortisol/17-OHP ratio, reflecting CYP21A2 activity, was significantly lower. In the mineralocorticoid pathway, 11-deoxycorticosterone, dehydrocorticosterone, and aldosterone were decreased, while in the androgen pathway dehydroepiandrosterone (DHEA) was increased. In the subgroup with genomic analysis, patients carrying only one C4B copy displayed reduced CYP21A2 activity, as reflected by a lower S/17-OHP ratio, and correlations were observed between C4 copy number, 17-OHP levels, and the S/17-OHP ratio.

**Conclusion:** Men with CSCR present systemic dysregulation of glucocorticoid, mineralocorticoid and androgen pathways, linked to impaired CYP21A2 activity. The correlation between low C4B copy number and altered steroid metabolism suggests a role of the RCCX module in CSCR susceptibility, warranting further genetic and functional investigations

## Introduction

Central Serous Chorioretinopathy (CSCR) is a retinal disease that predominantly affects males of mid-adulthood. It is characterized by the spontaneous separation of the neural retina from the retinal pigment epithelium (RPE), resulting from a disruption of the outer retinal barrier. In approximately one-third of the patient population, the disease manifests as bilateral, recurrent serous retinal detachments and is associated with blinding complications, including degenerative epitheliopathy and macular neovascularization (MNV). This renders CSCR the fourth retinal cause of visual impairment in the young population.^1–3^.

CSCR is a multifactorial disease with anatomical, genetic, environmental and exogenous risk factors ^4^. Anatomical factors including short eye (hyperopia) and thickened choroid with dilated vessels and abnormal regulation of the choroidal blood flow (pachychoroid) have been identified as predisposing phenotype ^5–7^ that precedes the occurrence of serous retinal detachments. Various genetic factors have been recognized as contributing elements to the development of CSCR. Specifically, polymorphisms in the vasointestinal peptide receptor 2 (VIPR2) gene have been associated with variations in choroidal thickness^8^. Polymorphism in NR3C2 (encoding the mineralocorticoid receptor) conferring greater hypothalamic pituitary adrenal (HPA)-axis reactivity and CRF (cortisol releasing factor) hypoactivity endophenotype ^9–11^ is associated with CSCR. An insertion variant in CRH (corticotropin releasing hormone) gene, that reduces its expression, was also recently associated with CSCR in the Chinese population ^12^. Polymorphism in proteins that regulate or participate to complement pathway activation, at risk for age related macular degeneration, are rather protecting for CSCR ^13^. In addition, genomic copy variation number in complement factor 4 (C4), and more specifically higher copy number of C4B has been shown to protect against chronic and severe forms of the disease ^14^.

Circadian rhythm disruption, shift work, sleep disorders and apnea have been demonstrated to be associated with CSCR^4,15,16^. However, the prevailing consensus among researchers is that the most significant risk factor is the systemic or locoregional intake of glucocorticoids (GCs), even weeks or months before the disease is detected ^17–19^. Since high doses of intraocular GCs, which do not diffuse in the circulation, are not associated with risk, this suggests that it is the systemic exposure and not the ocular exposure to GCs that can trigger and/or aggravate CSCR^20^. Conversely, the association between endogenous hypercortisolism and CSCRC ^21,22^ remains ambiguous. Hair cortisol levels, which most precisely reflect cortisol exposure in the months preceding a CSCR episode, did not demonstrate elevated levels^23^. Finally, in the ocular media of CSCR patients, lower endogenous glucocorticoid levels were measured in comparison to those observed in the control group ^24^. Therefore, the following hypothesis was formulated: a subclinical brake of the HPA axis, in response to systemic GCs intake^25^ and / or to genetic, epigenetic or environmental factors that disrupt corticoid metabolism, rather than corticoid excess could favor CSCR^20^.

Whether patients with CSCR present endogenous corticoid excess or insufficiency or any corticoid metabolism impairment cannot be evaluated by the measurement of serum cortisol, that is highly influenced by stress, particularly in CSCR patients, known to have a high stress sensitivity, exacerbated by their visual disturbance ^26,27^. Indeed, exploration of corticoid metabolism requires the analysis of the full steroidome which provides a comprehensive overview of the corticoids produced by the cortex of the adrenal glands through the transformation of cholesterol through successive enzymatic reactions ^28,29^ [Figure 1]. In the zona fasciculata of the adrenal cortex, the production of cortisol (F) is facilitated by P450c17 (CYP17), a 17α-hydroxylase/17,20 lyase. This enzyme converts pregnenolone to 17α-hydroxypregnenolone, which is subsequently converted into 17α-hydroxyprogesterone (17-OHP) by 3β-hydroxysteroid dehydrogenase (3HSD). The 21-hydroxylation of 17-OHP is subsequently facilitated by the steroid 21-hydroxylase P450c21 (CYP21), resulting in the formation of 11-deoxycortisol (S). This is then converted to cortisol by 11β-hydroxylase P450c11 (CYP11B1) within the mitochondria. In the zona glomerulosa, the P450c21 enzyme also catalyzes the conversion of progesterone into 11-deoxycorticosterone (DOC).

**Figure 1.**
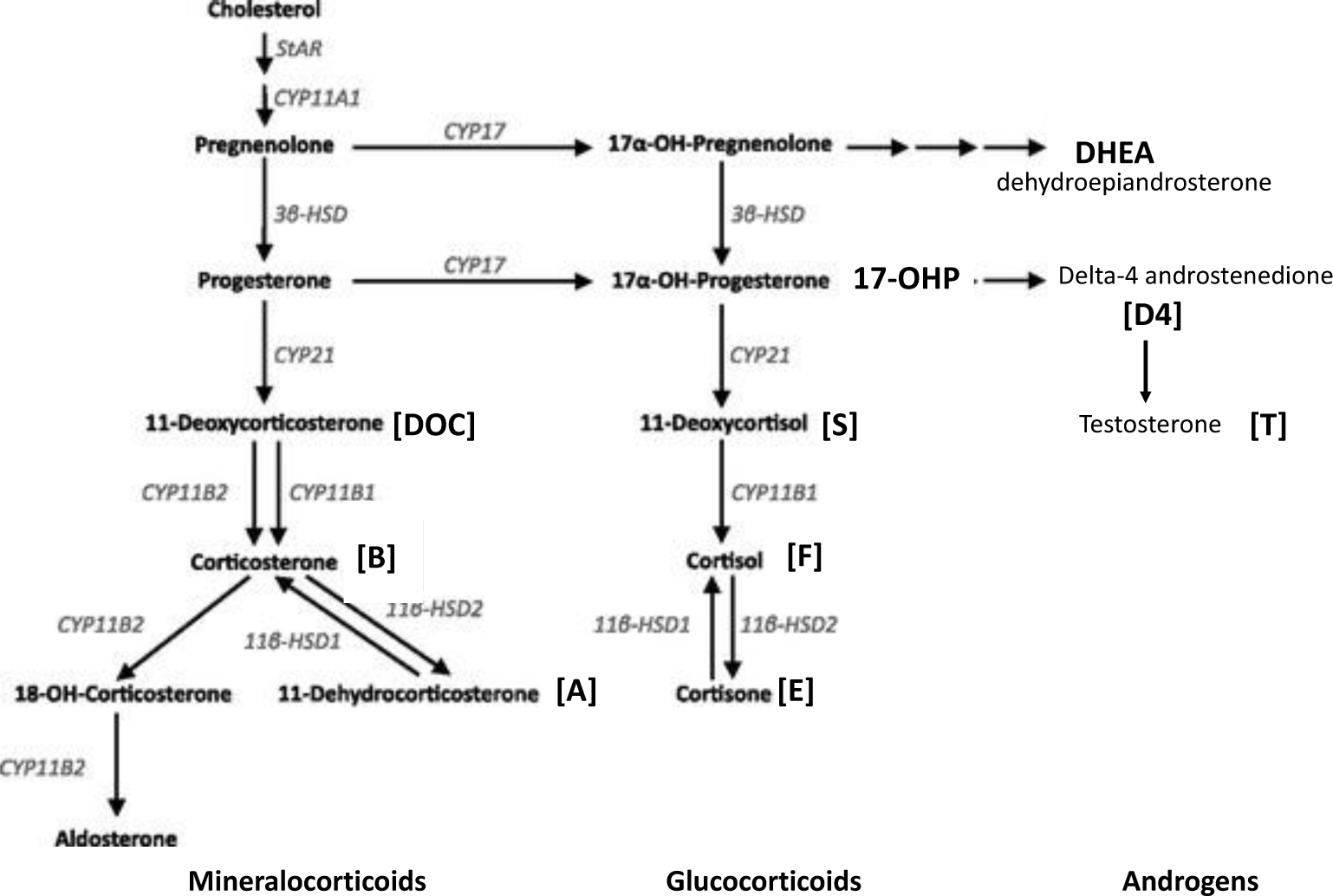
Adrenal metabolism of corticoids

Aldosterone, the most potent mineralocorticoid, is produced by the 11β-hydroxylation of DOC to corticosterone (B). The final three steps of aldosterone synthesis are catalyzed by the mitochondrial enzyme P450c11AS (CYP11B2). Within the zona reticularis, P450c17 catalyzes the conversion of 17α-hydroxypregnenolone to the androgen dehydrogenase androsterone (DHEA) [Figure 1]. The most recent liquid chromatography coupled to tandem mass spectrometry (LC-MS/MS) have been developed for the analysis of various corticoids. These methods provide information regarding the activity of distinct enzymatic reactions that occur in the adrenal glands^25,28^ and help identify enzymatic deficits ^30–32^.

The CY21 is encoded by the *CYP21A2* gene, which is located in the 6q21.3 region. In this region, the pseudogene *CYP21A1P* shares approximately 98% of homology with its coding region^33^. Both *CYP21* genes are aligned in tandem with *C4*, *SKT19* (encoding the serine/ threonine kinase 19) and the *TNX* (encoding tenascin X) genes (arranged as *SKT19-C4A-CYP21A1P-TNXA-STK19B-C4B-CYP21A2-TNXB*), located in the MHC class III region. They form the RCCX module that is a complex and multiallelic, tandem copy number variation (CNV)^34^, with a variable number of copies among individuals that controls gene expression, the protein functions, and the phenotypic traits, contributing to the genetic and phenotypic heterogeneity ^35,36^. A notable finding is that patients with high *C4B* copy number exhibit a substantially reduced of developing chronic CSCR and patients with less than 2 copies and particularly those with no copy of *C4B* are at high risk of presenting chronic CSCR ^37^. In contrast, patients with less than two copies of C4B, particularly those with no copies, demonstrate an elevated risk of developing chronic CSCR^37^. Concurrent with *C4* gene CNV is the CNV for *CYP21* gene^38^. According to the human genome assembly, *C4B* conjugates with *CYP21A2*, making *C4B* and *CYP21A2* susceptible to be lost together in the event of a deletion, like in congenital adrenal hyperplasia patients^39^. However, the potential relationship between the *C4* CNV and the steroidome of patients with CSCR remains uninvestigated, and the steroidome of patients with CSCR has not been thoroughly examined. In this study, we have conducted a comprehensive analysis of the steroidome in the serum of men diagnosed with CSCR, compared it with that of a matched group of men without CSCR, and explored the potential implications of these findings. To further explore the link between the complement C4 and corticoid metabolism, we also have analyzed the *C4A* and *C4B* copy number in a subset of the CSCR patients and have correlated them to corticoid metabolism markers.

### Subjects and methods

#### Ethics Statement

Caucasians men diagnosed with CSCR (n=45) from two cohorts, followed at the Jules Gonin Eye Hospital (Lausanne, Switzerland) and at the Ophtalmopôle Cochin hospital (Paris, France) were included. This research was conducted in compliance with the tenets of the Declaration of Helsinki and was approved by review boards in Switzerland (CER-VD Eyeomics 340/15) and in France (CPP Ile de France 1, C16-09 N°DC-2016-2620 and DC-2012-1514). Written informed consent was obtained for each patient for clinical data analysis and biological material analysis.

#### Blood collection

The steroidome was analyzed only in men serum to limit sex hormones variations due to cycles in women and because CSCR is more frequent in men. Serum from control men subjects were obtained from the blood French Bank (“Banque Française du Sang, BFS”) under an agreement between BFS and Inserm. Blood was prospectively collected from 49 healthy men donors who had no previous history of ocular or systemic diseases and were matched for age with CSCR patients. Patients and controls were sampled between 9am and 3pm. Serum were prepared and stored at −80°c in the biobank under similar conditions for CSCR and control individuals.

DNA from CSCR patients was prepared as previously described and stored at −80°c until analysis (DNA isolation in CSCR: https://pubmed.ncbi.nlm.nih.gov/34490249/).

#### Ocular phenotyping

Diagnosis criteria for CSCR were defined on history and on multimodal imaging including spectral-domain optical coherence tomography (SD-OCT, Spectralis, Heidelberg Engineering, Heidelberg, Germany), blue-light fundus autofluorescence (BAF) imaging (Spectralis) and, fluorescein and ICG angiography (Spectralis) according to the CSCR study group classification as previously described ^40^. Patients were classified as chronic or “complex” CSCR based on the presence of an underlying epitheliopathy characterized on blue autofluorescence and fluorescein angiography (total cumulated area >2 disk diameters), or they were classified as acute/recurrent or “simple” cases if there was no epitheliopathy. Patients with any other ocular disease such as age-related macular degeneration (characterized by the presence of drusen), diabetic retinopathy, retinal vein occlusion, high myopia >-6 diopters were not included in this analysis.

#### Steroidome analysis

Steroid concentrations in serum samples were measured by liquid chromatography coupled to tandem mass spectrometry, as previously reported ^24,28^. Briefly, steroids were purified from 200 μL of serum sample spiked with a mix of deuterated internal standards using a protein precipitation procedure followed by a solid phase extraction step (OASIS, Waters, Guyancourt, France). Aldosterone (ALDO), corticosterone (B), 11-dehydrocorticosterone (A), 11-deoxycorticosterone (DOC), 17-hydroxyprogesterone (17-OHP), 11-deoxycortisol (S), cortisol (F), cortisone (E), androstenedione (Δ4A), testosterone (T) and dehydroepiandrosterone (DHEA) were resolved using a BEH C18 column with a methanol-ammonium formate gradient during fourteen minutes. Detection was performed on a Xevo TQS tandem mass spectrometer (Waters, Paris, France) equipped with an electrospray operating in positive mode. Quantification was performed in the multiple reaction monitoring mode. The calibration curve samples were prepared in charcoal-treated serum. The data were fit to a linear least square regression curve with a weighing index of 1/*x*. Calibration ranges, limit of quantification (LOQ) and between-run coefficients of variation (%) for individual steroids in both media are shown in Table 1. The enzymatic activity index of P450c21 biosynthesis step was estimated by the 11-deoxycortisol to 17-hydroxyprogesterone ratio (S/17-OHP).

**Table 1.** Calibration ranges, limit of quantification (LOQ) and between-run coefficients of variation (%) for individual steroids in serum. B: corticosterone, A: 11-dehydrocorticosterone, DOC: 11-desoxycorticosterone, F: cortisol, E: cortisone, S: 11-desoxycortisol, 17-OHP: 17-hydroxyprogesterone, Δ4A: delta4-androstenedione, T: testosterone, DHEA: dehydroepiandosterone.

| Analyte | Calibration range (ng/ml) | LOQ (ng/ml) | Between-day CV (%) |
| --- | --- | --- | --- |
| Aldo | 0.022-2.7770 | 0.015 | 6.31 |
| B | 0.529-45.400 | 0.040 | 4.24 |
| A | 0.100-2.0 | 0.040 | 9.31 |
| DOC | 0.044-3.150 | 0.020 | 11.05 |
| F | 10.1-278.0 | 0.060 | 3.92 |
| E | 1.01-20.50 | 0.050 | 8.07 |
| S | 0.105-14.1 | 0.040 | 6.48 |
| 17-OHP | 0.094-14.200 | 0.040 | 8.12 |
| DHEA | 0.056-11.7 | 0.025 | 5.89 |
| T | 0.056-11.7 | 0.025 | 5.89 |

### Genomic C4 CNV evaluation

The copy numbers of C4 genes were measured by quantitative PCR according to the manufacturer’s protocol (Applied Biosystems, Thermo Fisher Scientific). Each 20 μl qPCR reaction included 20 ng of genomic DNA, 10μl of 2X qPCR Master Mix, 1μl of 20X VIC dye-based TaqMan Copy Number Reference Assay for RNase P, and 1 μl of 20X MGB dye-based TaqMan Copy Number Assay for either C4A (Hs07226349_cn) or C4B (Hs07226350_cn). All assays were run in triplicate on a QuantStudio 3 instrument (Applied Biosystems, Thermo Fisher Scientific), with no-template controls (NTCs) included for a quality control. The cycling conditions comprised an initial 10-minute step at 95°C for denaturation and enzyme activation, followed by 40 cycles of 15 seconds at 95°C and 60 seconds at 60°C. Copy number calculations were analyzed using QuantStudio Design and Analysis 2 (Applied Biosystems, Thermo Fisher Scientific), using the ΔΔCt method normalized to the reference RNase P.

#### Statistical analysis

Quantitative data was tested for normality with the Shapiro-Wilks tests. They are reported as means with standard deviations (SD) when normally distributed and medians with interquartile ranges (IQR) when non-normally distributed. Data were compared between the two groups using unpaired t test with Welch’s correction for normal distributions and using Mann-Whitney U-tests for non-normal distributions. Correlation between two quantitative variables was evaluated using Spearman test. p-values < 0.05 were considered statistically significant. Statistical analyzes were performed using GraphPad Prism (version 9.1.2 (226), GraphPad Software, LLC). This study has been conducted following another study in which we have measured the steroidome in control individuals with different age and sex, using the same analytical method^24^, allowing to estimate the number of men needed to find a difference between a control group and a diseased group using the biostatgv.sentiweb.fr software.

## Results

### CSCR population data

Caucasian men (n=45) with CSCR were included in the steroidome analysis study. The mean age was 53.6±11.9 years [32-70]. Regarding known risk factors, 36% had a history of GCs intake (6 oral, 3 inhaled, 4 intraarticular, 1 oral and inhaled, 2 dermal administration) but none were under steroid treatment at the sampling time. Recent stressful psychological event (in a professional or personal setting) was reported by 40% of the patients, 10% had hypertension, 11% presented sleep apnea, 27% suffered from chronic allergy and 11% reported shift work. 32 patients had complex CSCR, amongst which 7 had MNV and 13 patients had simple acute CSCR. The mean subfoveal choroidal thickness was 487±94 µm[300-721µm]. The mean age of the 49 healthy men was 52.6 ± 11.48 years and did not differ from the mean age of the CSCR men (p=0.67).

### Steroidome in CSCR patients

In the glucocorticoid pathway, the mean level of 17-OHP was significantly higher in patients with CSCR as compared to controls (0.85±0.40 vs 0.6±0.20, unpaired t test, p=0.005) while the level of 11-deoxycortisol (S) did not differ in both groups (0.20 ±0.174 vs 0.21 ±0.10, MW test p=0.1) (Figure 2A, B). The index of CYP21 activity (S/17-OHP) was significantly lower in CSCR patients than in controls (0.36 ±0.55 vs 0.41±0.29, mean ranks 38.59 vs52.94; p= 0.009) (Figure 2C). There was no significant difference in the cortisol (F) level between CSCR and controls (98.2 ±35.9 ng/ml vs 104.8 ± 28.3, MW test p=0.17) but the level of cortisone (E) was significantly lower in CSCR patients (18.4 ± 4.6 vs 41.2 ± 4.6 ng/ml, p=0.0004, unpaired t test) (Figure 2D, E).

**Figure 2.**
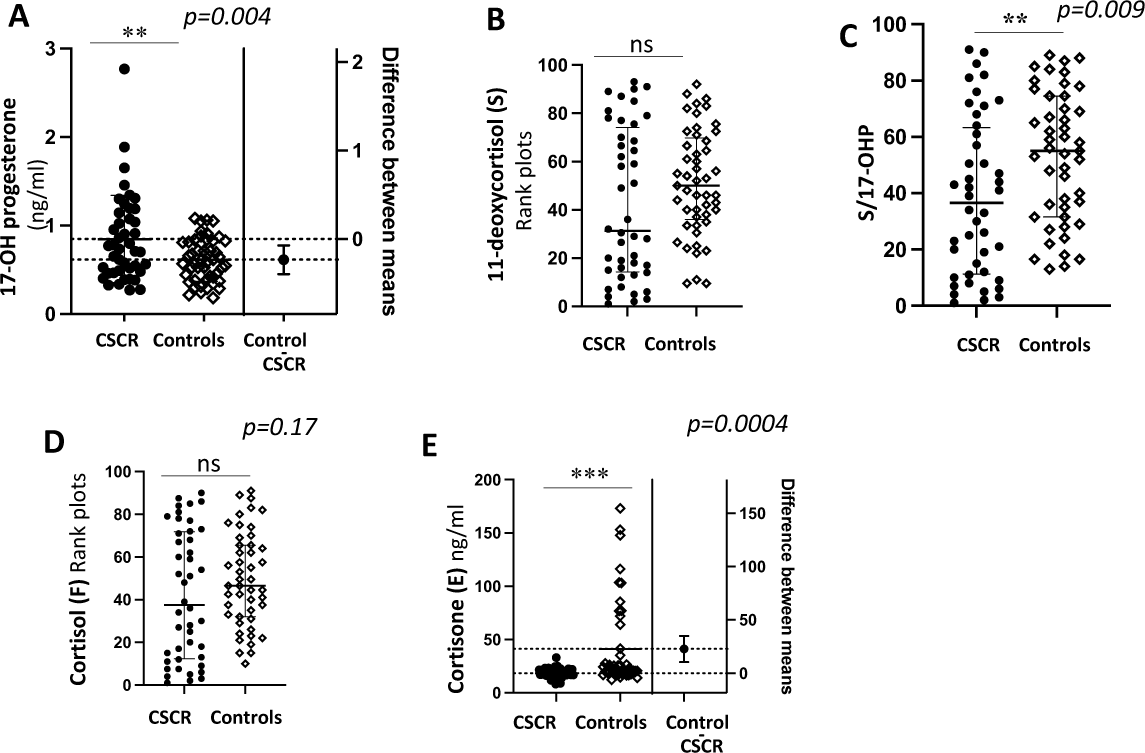
Glucocorticoids pathway in men with CSCR. Serum concentrations (in ng/ml) of 17-OHP [**A**], 11-deoxycortisol (S) [**B**], ratio S/17-OHP [**C**], cortisol (F) [**D**], cortisone (E) [**E**]. Values are reported as means ± SD when normally distributed as medians with interquartile ranges when non-normally distributed. ns: non-significant.

In the mineralocorticoids pathway, 11-deoxycorticosterone (DOC) was reduced in CSCR group as compared to the control group (0.025 ± 0.016 vs 0.06 ± 0.053, unpaired t-test, p<0.0001).

Corticosterone (B) was similar in both groups (1.89±1.44 vs 1.69±1.38, MW test p=0.43) but its 11-oxidized derivative, 11-dehydrocorticosterone (A) was significantly lower in CSCR patients as compared to controls, 1.33± 1.15 vs 1.60 ± 0.75, MW test p=0.009) (Figure 3A,B, C). Finally, aldosterone was also significantly lower in the CSCR group as compared to controls (0.05 ±0.05 vs 0.084±0.056 ng/ml, MW test p=0.024) (Figure 3D).

**Figure 3.**
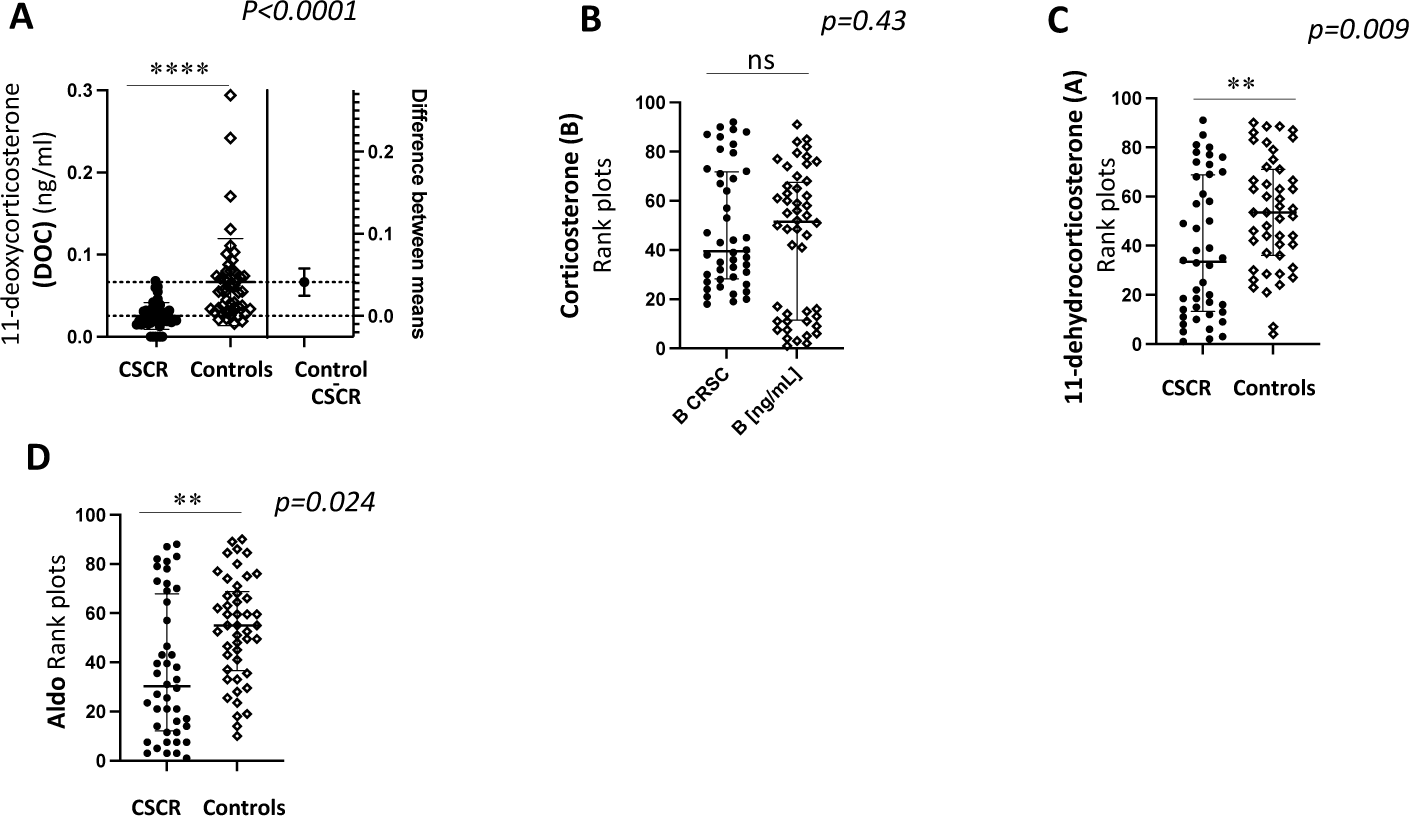
Mineralocorticoid pathway in men with CSCR Serum concentrations (in ng/ml) of 11-deoxycorticosterone (DOC) **[A**], corticosterone (B) [**B**], 11-dehydrocorticosterone (A) [C], aldosterone (Aldo) [**D**]. Values are reported as means ± SD when normally distributed as medians with interquartile ranges when non-normally distributed. ns: non-significant.

In the androgen pathway, delta-4 androstenedione was similar in both groups (0.68±0.26 vs 0.71±0.32, unpaired t test, p=0.63) but the level of DHEA was significantly increased in CSCR as compared to controls (11.07 ±6.62 vs 5.65 ± 4.8, p<0.0001) (Figure 4A, B). Finally, the level of testosterone was not different between the two groups (4.62 ± 1.67 vs 5.13 ±1.73, unpaired t test, p=0.16) (Figure 4 C).

**Figure 4.**
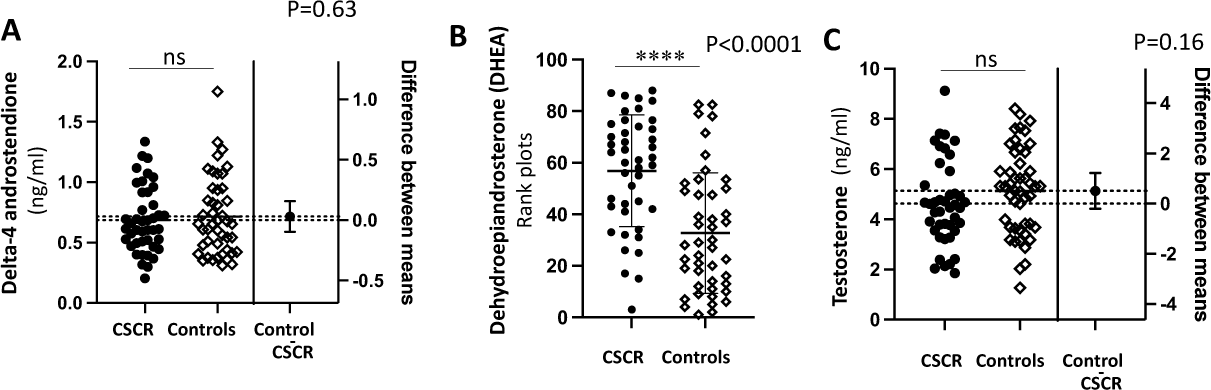
Androgen pathway in men with CSCR Serum concentrations (in ng/ml) of delta-4 androstenedione [**A**], dehydroepiandrosterone (DHEA) [**B**] and testosterone [**C**]. Values are reported as means ± SD when normally distributed as medians with interquartile ranges when non-normally distributed. ns: non-significant.

There was no significant difference in any of the measured corticoids between CSCR patients suffering from acute/ simple or chronic/ complex form of the disease (not shown).

Figure 5 recapitulates the metabolites, which levels are different in CSCR as compared to controls, indicating possible decreased activity of CYP21.

**Figure 5.**
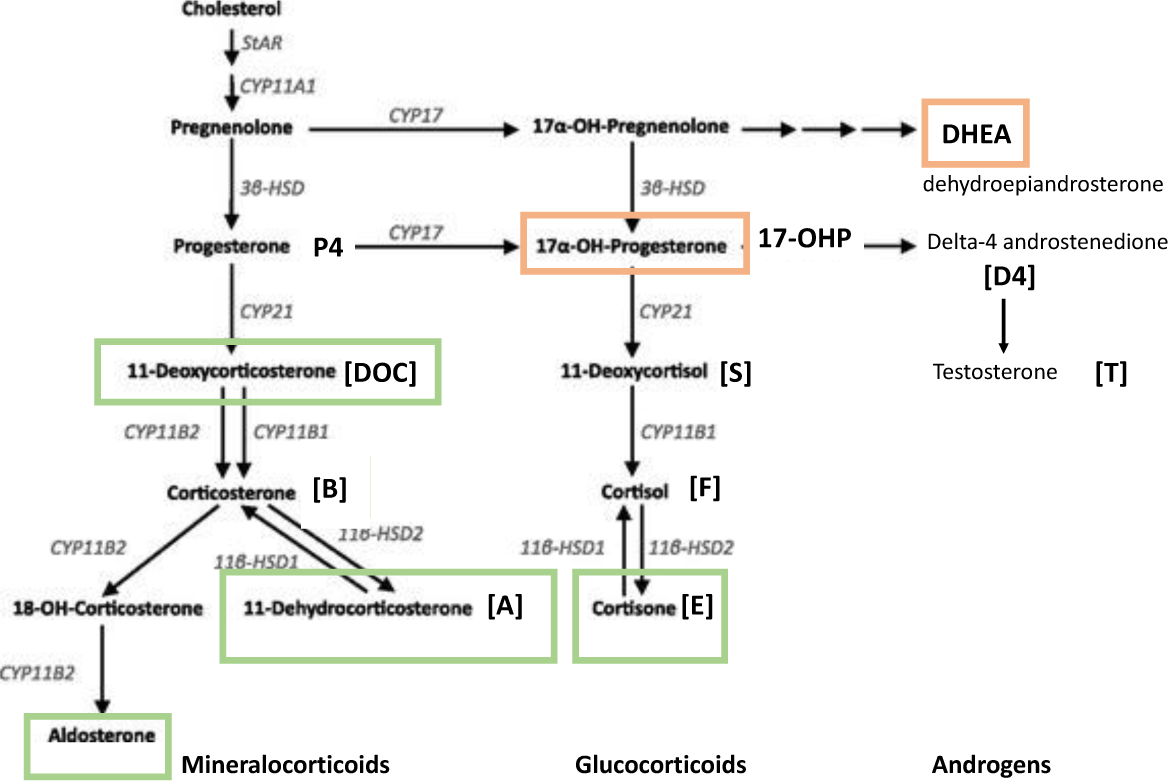
Steroidome of patients with CSCR Metabolites lower (in green) or higher (in red) in men with CSCR, suggesting reduced CYP21 activity.

### Correlation between genomic C4 copy number and corticoid metabolism in CSCR

In a subgroup of 20 patients with chronic/complex CSCR who provided consent for genetic analysis, the genomic C4 copy numbers were quantified, and their correlation to 17-OHP and the metabolic ratio S/17-OHP ratio, used as an indicator of CYP21 activity, was determined.

The mean number of total C4 copies was 4.2 ± 1.2 [2.1-6.8], with 2.7 ± 0.9 [1-5] copies of C4A and a low number of copies of C4B, as patients with CSCR had either 1 or 2 copies of C4B (Figure 6A). The observed variation in the number of copies of total C4 was predominantly associated with the variation in C4A CNV.

**Figure 6.**
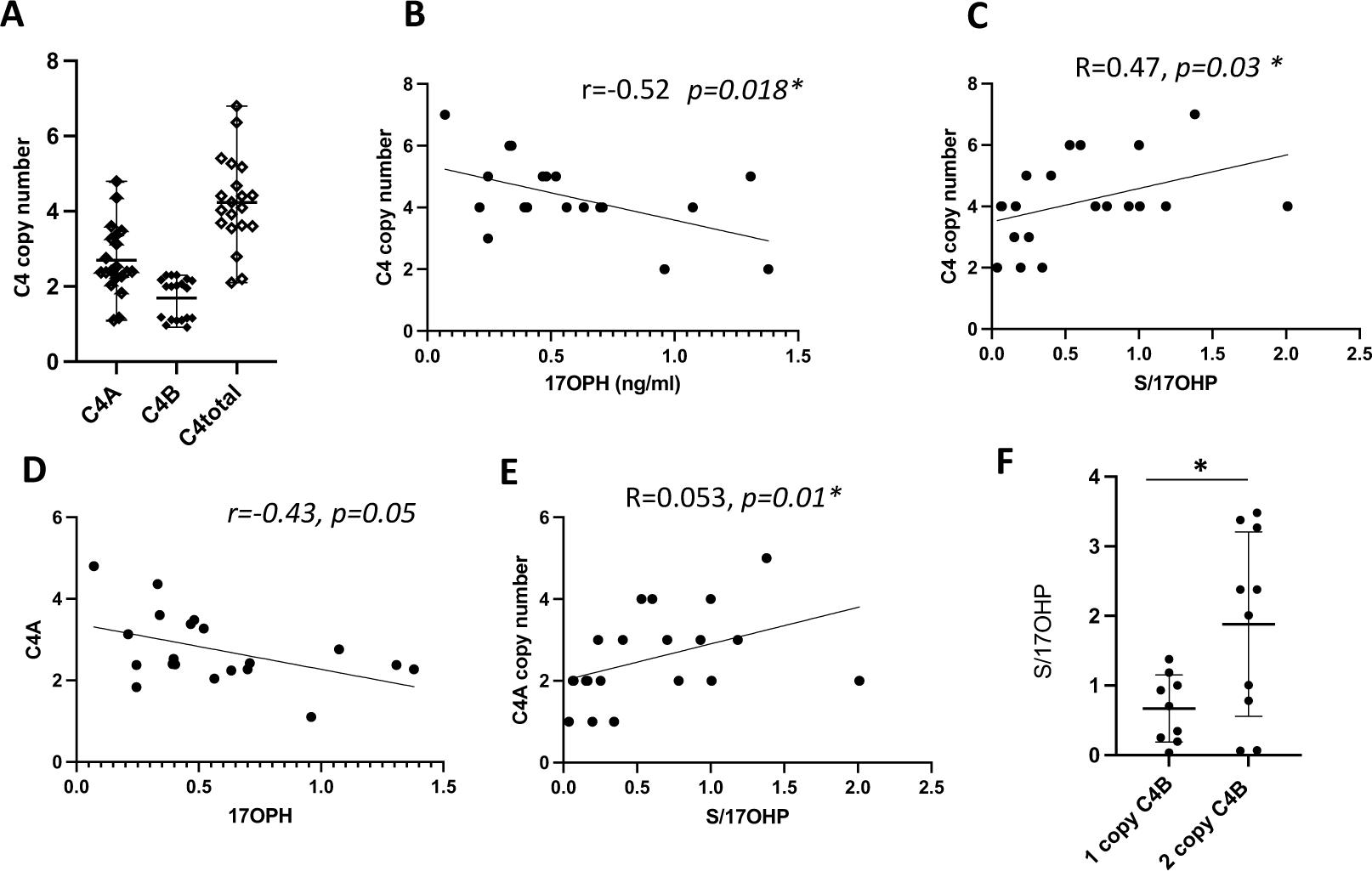
Genomic C4 copy number and correlation with corticoids in men with CSCR A: Number of C4A, C4B and total C4 in men with chronic CSCR, **B:** Correlation between C4 copy number and 17OHP, **C:** Correlation between C4 copy number and S/17-OHP ratio, **D:** Correlation between C4A copy number and 17-OHP, **E:** Correlation between C4A copy number and S/17-OHP ratio, **F**: S/17-OHP ratio in patients with chronic CSCR and either one or 2 copies of C4B.

A correlation was identified between genomic C4 copy number and the levels of 17-OHP (p=0.018) and the ratio S/17OHP (p=0.03) (Figure 6B, C). A significant correlation was also observed between C4A copy and the S/17OHP metabolic ratio (p=0.01) (Figure 6E). The S/17-OHP ratio was significantly lower in the group of patients with one copy of C4B as compared to those with two copies (0.66 ± 0.48 vs. 1.88 ± 1.32, p = 0.01).

## Discussion

The link between glucocorticoids and CRSC is complex and merits further exploration. The use of exogenous glucocorticoids by the extraocular route has been recognized for many years as a risk factor for the disease onset and severity^17–19^. However, if exposure of ocular tissues to high doses of glucocorticoids could cause CRSC in predisposed individuals, we would have expected to see a dramatic increase in CRSC in patients treated with intravitreal implants that deliver high doses of corticosteroids to the pathologically affected tissues (RPE and choroid), which was not the case. The other surprising fact is the temporal relationship between GCs use and disease, with a possible delay of several weeks or months. In this context, it has previously been posited that the disease-promoting factor may be the inadequate braking of the hypothalamic-pituitary axis, leading to corticoid deficiency, rather than corticoid excess^25,20^.

Psychological stress, which is also a trigger for disease or episodes of disease activity, has long been thought to act by increasing endogenous glucocorticoids. However, recent studies have highlighted a more complex psychopathology in patients with CRSC, with a predisposition to maladaptive personality, depression or post-traumatic syndromes ^26,26,41,42^, which may be associated with both cortisol elevation or deficiency^43,44^. Furthermore, numerous studies have examined the levels of circulating cortisol in patients with CRSC. However, these studies have predominantly compared these levels with those observed in unaffected controls. This is an important limitation, as CRSC itself is associated with stress, which is related to visual impairment. A subsequent study compared cortisol levels in patients with CRSC with those in patients with other types of retinal detachment and found no significant differences^45^. Moreover, it is now widely acknowledged that to thoroughly examine the metabolism of corticoids, that is, the steroidome, a comprehensive perspective on the successive enzymatic reactions leading to their production is imperative. This approach, founded on the principles of mass spectrometry, is regarded as the gold standard for such investigations^29^. The targeted liquid chromatography-tandem mass spectrometry (LC-MS/MS) analysis method was employed to assess the ocular steroidome and its relationship with systemic corticoids. The findings revealed a decrease in gluco- and mineralocorticoid metabolites within the aqueous humor of patients diagnosed with chronic inactive CSCR^24^. In the present study, we measured lower serum levels of cortisone, 11-deoxycorticosterone, 11 dehydrocorticosterone and aldosterone in men with CSCR than in age-matched male controls. This results agrees with genetic studies showing that the *NR3C2* and the *CRH* polymorphisms ^9,12^ which confer risks for CSCR, are associated with reduced glucocorticoids production. In addition, in CSCR patients, circulating lipocalin 2 ^46^ and pentraxin 3 ^47(p3)^, two markers of glucocorticoid pathway activation ^48,49^ were lower than in unaffected individuals and the metabolome of men with CSCR did not indicate glucocorticoid excess ^50^. Altogether, these different types of information converge towards the conclusion that it is not glucocorticoid excess that favor CSCR but a more complex dysmetabolism of endogenous corticoids leading to insufficient ocular glucocorticoid pathway activation.

In the present study, the cortisol levels did not differ between CSCR and controls, but this result cannot be fully interpreted because we did not strictly control the time of sampling and because cortisol is sensitive to psychological stress resulting from CSCR-induced visual changes. Nevertheless, the steroidome shows a significant reduction of both gluco and mineralocorticoid metabolites, confirming that some men with CSCR produce insufficient adrenal corticoids.

The mechanistic link between corticoid dysmetabolism and CRSC remains to be understood. In a rodent model, the reduction in adrenal corticoid production, following a braking of the HPA axis, caused a decrease in ocular glucocorticoids, similar to what we have measured in the aqueous humor of patients with inactive CSCRc^24^, together with a hyperactivation of the ocular mineralocorticoid (MR) pathway^25^. In the posterior segment of the eye, as in many tissues, MR pathway overactivation is pathogenic^51,52^. Transgenic rodents overexpressing the human MR or chronic exposure to systemic aldosterone (which is a MR ligand), both induced a pachychoroid phenotypec^53,54^ and a choroidal neuropathyc^55^, which predispose to CSCR, although no animal model fully recapitulates the human phenotype of CSCR.

The CSCR steroidome exhibited an increase in 17-OHP and DHEA, suggesting that some CSCR men may experience partial CYP21 activity insufficiency, manifesting as an asymptomatic condition, a phenomenon frequently observed in men with non-classical congenital adrenal hyperplasiac^31,56^. In certain cases where 17-OHP levels were not elevated, adrenal insufficiency may have been attributable to exogenous corticosteroid therapy, chronic stress, or environmental factors, such as exposure to endocrine-disrupting chemicals, as suggested by the metabolomic signature in CSCR ^57,58^. The corticoid metabolism in patients with CSCR is likely a product of a combination of genetic and environmental factors.

The *CYP21* gene resides in the RCCX module, that also contains the genes encoding complement component 4. RCCX is a complex, multiallelic and tandem CNV (copy number variation) located in the major histocompatibility complex (MHC) class III region^33,36,59^. It consists of 1–3 tandem repeats of a DNA segment on one chromosome, each DNA segment encoding 2 complete genes, *C4* and CYP21, in equal copy numbers^34^.

According to the findings of previous studies, the risk of chronic CSCR is contingent upon *C4B* genomic copy numbers, with the absence of *C4B* copy resulting in elevated risk ^37^. In our study, men with chronic CSCR were found to have either one or two C4B copies. The S/17OHP ratio, which reflects the activity of CYP21, is reduced in patients with CSCR, and it correlates with the number of C4 copy numbers. This phenomenon can be explained by the fact that an intrinsic part of the *CYP21* gene promoter is located in the *C4* gene.^60^ A low number of C4 copy numbers would result in lower activity of CYP21.

This study is not without its limitations, including the size of the studied population. However, it is noteworthy that this is the largest CSCR cohort with systemic steroidome analysis. To validate these preliminary observations, additional studies are necessary to confirm the findings. A notable limitation of the study is the absence of a comprehensive genetic analysis of the RCCX module in CSCR men. This was not the objective of the study, and the results indicate, particularly in patients with high 17OHP, the necessity for a thorough genetic study. Should the hypothesis be substantiated, individuals afflicted with subclinical nonclassical adrenal hyperplasia may reap benefits from a personalized hormonal therapy regimen..

In conclusion, the systemic steroidome analysis in men with CSCR showed an alteration of corticoid metabolism involving the gluco-, mineralocorticoid and androgen pathways, that could be related, at least for part of the patients, to a reduced activity of CYP21. The low CYP21 activity that correlates to low C4 copy number could indicate a link between chronic CSCR and the RCCX module, which should be further explored by in depth by genetic studies.

## Data Availability

All data produced in the present study are available upon reasonable request to the authors

## Acknowledgements

ANR MR-A-MD (PCRE-2020-20-CE17-0034) and ANR NEUROCOR (PCR-2023) for the financial support. We thank The Abraham J. & Phyllis Katz Foundation for their financial support for conducting research.

## Ethics Statement

This research was conducted in compliance with the tenets of the Declaration of Helsinki and was approved by review boards in Switzerland (CER-VD Eyeomics 340/15) and in France (CPP Ile de France 1, C16-09 N°DC-2016-2620 and DC-2012-1514). Written informed consent was obtained for each patient for clinical data analysis and biological material analysis.

## Author Contributions

Conceptualization: FBC, EB, CR, EP Methodology: MZ, JHH, MQ, AD, YA Formal Analysis: MZ, JHH, MQ Investigation: MZ, EB, AM, LK, MU Resources: CR, EP, FBC, YA Writing – Original Draft: MZ, JHH, FBC Writing – Review & Editing: All authors Supervision: FBC, EP, CR Funding Acquisition: FBC, EP, CR

